# Demonstrating the Utility of Instrumented Gait Analysis in the Treatment of Children with Cerebral Palsy

**DOI:** 10.1101/2023.10.13.23296949

**Authors:** Michael H. Schwartz, Andrew J. Ries, Andrew G. Georgiadis, Hans Kainz

## Abstract

**Background:** Instrumented gait analysis (IGA) has been around for a long time but has never been shown to be useful for improving patient outcomes. In this study we demonstrate the potential utility of IGA by showing that machine learning models are better able to estimate treatment outcomes when they include both IGA and clinical (CLI) features compared to when they include CLI features alone.

**Design:** We carried out a retrospective analysis of data from ambulatory children diagnosed with cerebral palsy who were seen at least twice at our gait analysis center. Individuals underwent a variety of treatments (including no treatment) between sequential gait analyses. We fit Bayesian Additive Regression Tree (BART) models that estimated outcomes for mean stance foot progression to demonstrate the approach. We built two models: one using CLI features only, and one using CLI and IGA features. We then compared the models’ performance in detail. We performed similar, but less detailed, analyses for a number of other outcomes. All results were based on independent test data from a 70%/30% training/testing split.

**Results:** The IGA model was more accurate than the CLI model for mean stance-phase foot progression outcomes (RMSE_IGA_ = 11^∘^, RMSE_CLI_ = 13^∘^) and explained more than 1.5× as much of the variance (R^2^*_IGA_* = .45, R^2^*_CLI_* = .28). The IGA model outperformed the CLI model for every level of treatment complexity, as measured by number of simultaneous surgeries. The IGA model also exhibited superior performance for estimating outcomes of mean stance-phase knee flexion, mean stance-phase ankle dorsiflexion, maximum swing-phase knee flexion, gait deviation index (GDI), and dimensionless speed.

**Interpretation:** The results show that IGA has the *potential* to be useful in the treatment planning process for ambulatory children diagnosed with cerebral palsy. We propose that the results of machine learning outcome estimators — including estimates of uncertainty — become the primary IGA tool utilized in the clinical process, complementing the standard medical practice of conducting a through patient history and physical exam, eliciting patient goals, reviewing relevant imaging data, and so on.

## 1. Background

### Instrumented Gait Analysis Past

> “I told you these were shadows of the things that have been,” said the Ghost. “That they are what they are, do not blame me!”

> — Charles Dickens, A Christmas Carol

Instrumented gait analysis (IGA) has been around for a long time but has never been shown to be useful for improving patient outcomes. By IGA we are referring to the instrumented aspects of a typical three-dimensional gait analysis evaluation. Specifically, kinematics, kinetics, and electromyographic signals. A full clinical gait evaluation generally also includes a complete physical examination, an observational video, review of patient birth, developmental, and treatment history, discussion of goals, functional limitations, and so on. But these data could be obtained in an ordinary clinic setting, and so they are not part of what we are calling IGA.

Examining our center’s historic trends of overall gait outcomes after major surgery reveals a picture of modest treatment effects that are stagnant and unpredictable (Figure 1). The use of IGA for three decades has not resulted in better or more consistent results. It could be argued that the utility of IGA was fully realized before 1994, but this is highly implausible considering all the improvements in equipment, advances in modeling, published scientific findings, and practical clinical experience that has accumulated during the interim.

**Figure 1:**
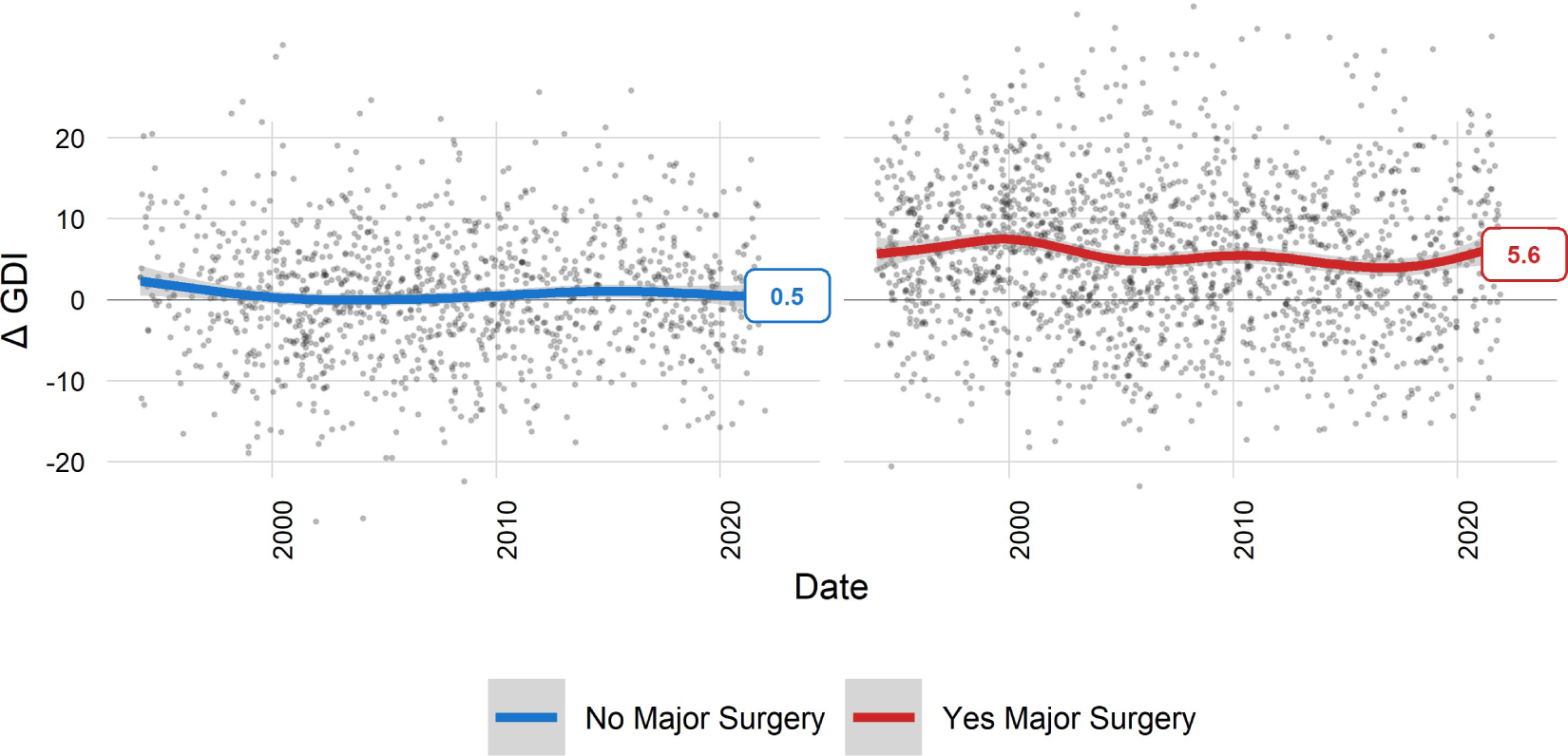
Trends in short-term outcomes for the Gait Deviation Index (GDI) show modest treatment effects (∼ 5 pts) that are stagnant and unpredictable. The LOESS smoothing spline (span = .7) reflects the moving average.

### Instrumented Gait Analysis Present

> “This boy is Ignorance. This girl is Want. Beware them both, and all of their degree, but most of all beware this boy, for on his brow I see that written which is Doom, unless the writing be erased. Deny it!” cried the Spirit, stretching out its hand towards the city. “Slander those who tell it ye. Admit it for your factious purposes, and make it worse. And bide the end!”

> — Charles Dickens, A Christmas Carol

From a theoretical perspective, the utility of IGA seems plausible. The output of IGA produces measurements of the underlying gait biomechanics and where they deviate from those of typically developing individuals. Thus, IGA identifies biomechanical impairments. The argument for the utility of IGA then goes something like…

Step One: Measure the biomechanical impairments present in the child’s gait
…
Step Three: Assign treatments that will improve some aspect of gait, mobility, or quality-of-life

Alert readers will notice that there is a step missing. We will call this gap between the first and third steps “Step Two”. Step Two is the process for deciding what treatments (if any) will achieve the desired outcome. In a three-step process, Step Two is often the trickiest, as suggested by Harris (Figure 2).

**Figure 2:**
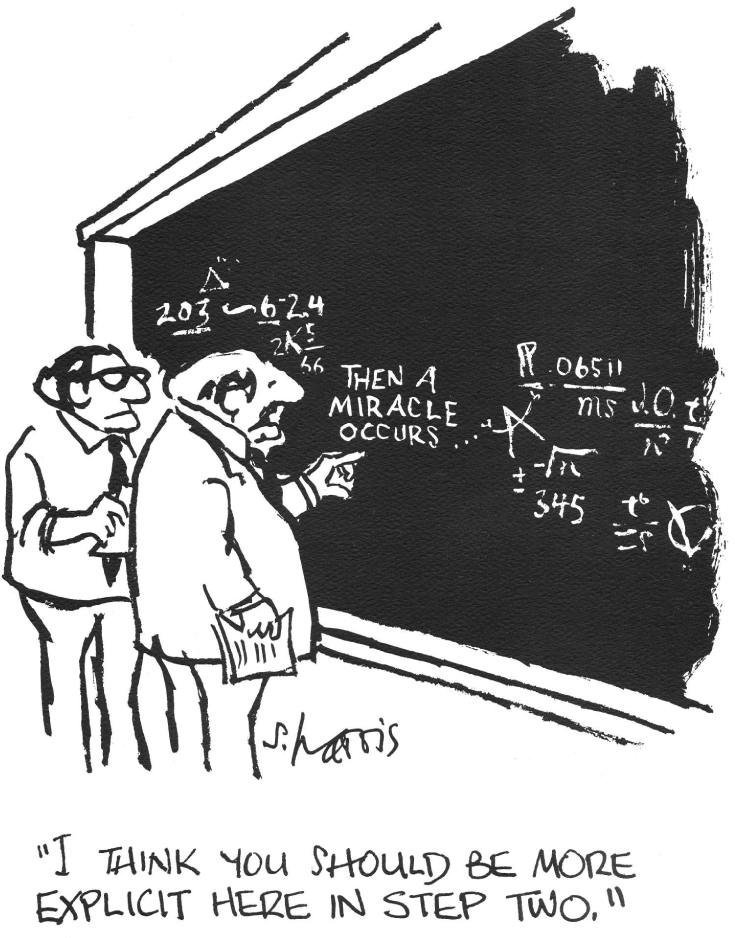
The notoriously difficult Step Two (Copyrighted artwork ScienceCartoonsPlus.com. All materials used with permission.)

If Step Two is missing, then how is it that children who undergo IGA end up with prescribed treatments? While there is variation in the process, treatment assignment (prescription) is usually decided by a group of people staring at hundreds of graphs, reading hundreds of clinical measurements, watching videos, reading dozens of paragraphs of patient history, viewing dozens of medical images, and often reminiscing about previous patients who may have presented with similar clinical and biomechanical profiles, may have undergone some treatment or another, and may have had some outcome — though all of these details are clouded by fallible human recall [1]. The interpreting team may also discuss published studies related to this surgery or that therapy — but these discussions are also generally based on memories of studies the discussants happen to have read. In short — the process is an earnest but unstructured and unscientific meeting of the minds, based on subjective impressions and sparse recollections. It is not, in any way, what we would call an evidence-based process.

In defense of the *status quo* — there is no viable alternative. Some centers add elements of evidence where possible. For example, at our center a standard IGA report includes estimated hamstrings length as part of the indications for hamstring lengthening surgery. We carefully follow the proposed guidelines of Arnold, et al. [2]. Perhaps as a result of our close adherence to this explicit, evidence-based guidance, and despite the fact that our center tends to be extremely conservative in administering hamstrings lengthening surgery (or maybe because of it?), we found that hamstrings lengthening was a top performer for causal treatment effects among all the surgeries we routinely administer [3].

This charming story about hamstrings lengthening suggests that IGA might be useful in assigning treatments that improve mobility in patients. But one confirmatory anecdote does not equate to scientific evidence. The ideal way to discover if IGA has value would be to conduct a randomized controlled trial (RCT). In this imagined RCT patients would be randomly assigned to undergo the instrumented aspects of IGA or not prior to treatment.

That is, one group would have pre-treatment CLI + IGA and the other would have only CLI. After enrolling a sufficient number of participants, the outcomes (and treatments) would be compared between the groups. If IGA has value, then there would be better outcomes for the group randomized to undergo CLI + IGA. Or, at a minimum, equal outcomes with fewer treatments. Nothing could be more straightforward. And, given that there is no evidence for the utility of IGA, there are no ethical obstacles to performing such a study.

The RCT described above is the ideal situation. But, reality rarely matches our expectations. Despite existing for nearly four decades, there are no RCTs examining the efficacy of IGA — reiterating that when we use the term IGA we are specifically referring to the instrumented bits, like kinematics, kinetics, and EMG. There are a number of reasons generally cited to explain this situation. Chief among these is that, at hospitals with active IGA centers, clinicians and patients generally believe IGA is valuable and are therefore resistant to eschewing it. A second challenge is that the heterogeneity of patients and treatments suggest the need for huge numbers of participants for an RCT to obtain reliable estimates of IGA’s effect.

In 2020, Wren and colleagues published an update to their 2011 systematic review examining the efficacy of gait analysis [4,5]. Between the two systematic reviews, the authors evaluated 11,696 articles, and found 2952 that met their inclusion criteria, 7 of which addressed individual patient outcomes [6–12]. Two of the articles were derived from a single RCT, while the others were either prospective cohort studies or retrospective analyses.

None of the studies directly addressed the value of IGA since they all failed to isolate IGA from the routine clinical portion (CLI) of a gait evaluation. The general design of the studies involved examining treatment outcomes stratified by adherence to recommendations derived from a gait evaluation. In other words, the studies compared (CLI + IGA + *adherence*) to (CLI + IGA - *adherence*). This isolates the role of adherence, but obviously fails to isoloate the role of IGA.

Some authors were clear about this shortcoming. For example, Lee et al. stated (emphasis added) “*The **<u>combination</u>** of a careful clinical assessment and gait analysis can produce better results in surgery for children with cerebral palsy*”. In their paper, Gough and Shortland asked “*Would it have been possible to define [patients who benefit from treatment] without gait analysis?*” They suggested IGA was needed by showing stronger associations between outcomes and IGA elements than between outcomes and CLI elements. This evidence is intriguing, but far from definitive. Surprisingly, many authors failed to address — or possibly even notice — that their studies did not isolate the role of IGA.

There were two studies derived from the single RCT that deserve additional attention, given the potential strength of randomization and the almost reflexive deference that the term RCT in a study title often engenders. Like the retrospective and cohort studies, the RCT examined the role of adherence to recommendations. The first of the two publications that followed the RCT found an effect of adherence on the Child Health Questionnaire and the upper extremity portion of the Pediatric Outcomes Data Collection Instrument. Considering that the study found no underlying differences in gait, both effects seem implausible.

Additionally, these outcomes did not match the primary outcomes pre-registered with clinicaltrials.gov (NCT00114075), which were (1) change in walking ability at 1 year and (2) change in walking efficiency at 1 year. The pre-registered secondary outcomes were (1) change in gross motor function at 1 year, (2) change in quality of life at 1 year, and (3) additional treatment needed at 1 year.

The next publication derived from the RCT reported on a “*secondary analysis*”. It examined the impact of adherence to recommendations on outcomes following a femoral derotation osteotomy (FDO). There was no effect in the primary outcome measure, as stated by the authors (emphasis added), “*Outcomes **<u>did not differ</u>** between the group which received a gait report … and the control group*”. The authors proceeded to identify a subgroup that did exhibit an effect. Thus, an effect of adherence to recommendations (not IGA) was found in a subgroup of a secondary analysis not described in the RCT pre-registration. *Post hoc* secondary and subgroup analyses are common, but run a substantial risk of artificially inflating statistical significance. This is true even in the absence of ill-intent. Thus, the conclusions that ensue from such analyses must be treated with a great deal of skepticism [13].

The seven studies identified by two systematic reviews tested the value of recommendations derived from complete gait evaluations (CLI + IGA). If we assume that the recommendations were partially influenced by IGA, then IGA can potentially be given credit for an undetermined portion of small effects, some of which were implausible, and others that were derived from unregistered secondary analyses of subgroups. Keep in mind that these effects were found in only seven papers out of 2952 (or 11,696 depending on how you count) published over the course of more than three decades since IGA’s inception. At this point we reiterate the fundamental claim of the present study: “*Instrumented gait analysis (IGA) has been around for a long time but has never been shown to be useful for improving patient outcomes.*”

### Instrumented Gait Analysis Yet to Come

> “Men’s courses will foreshadow certain ends, to which, if persevered in, they must lead,” said Scrooge. “But if the courses be departed from, the ends will change. Say it is thus with what you show me.”

> — Charles Dickens, A Christmas Carol

In this study we propose a new approach for testing the utility of IGA. We will test whether using data available only from IGA improves the precision and accuracy of machine learning (ML) models that predict outcome. If outcomes are more faithful, precise, or responsive using IGA data, then we conclude that there is potential value for IGA. Why only “*potential*” value? Because the value can only be realized if the data are used in a way that actually generates superior outcomes. That is, we need to define a proper Step Two. Our study proposes that Step Two is to use ML models to estimate patient outcomes from various treatments, then use these estimates to guide treatment assignment. In other words, our proposed IGA process is

Step One: Measure the biomechanical deviations present in the child’s gait

### Step Two: Use ML algorithms to estimate outcomes for various treatments

Step Three: Use the estimated outcomes (Step Two) to guide treatment assignment. That is, use the probability of positive outcomes in conjunction with other standard clinical practices (e.g., patient goals, risk assessment, etc.) to decide what treatments to render.

This approach has several advantages:

- *It is evidence-based*. The models use all available treatments and outcome information rather than only what can be recalled while reviewing the results of the IGA study.
- *It is transparent*. The models are open and available to clinicians and patients.
- *It is objective and reproducible*. Anyone who runs a patient’s data through the model will obtain the same estimated outcome every time.
- *It provides honest estimates of uncertainty*. The models produce rigorous distributions of possible outcomes. These distributions can be queried for a specific certainty (e.g., 90% prediction interval within which a patient’s outcome will be found 90% of the time). Clinicians and families can use this information in their decision-making process, taking into account their own risk tolerance.

In what follows we will describe our proposal in detail. In doing so, we will provide evidence that IGA has potential utility for treating children with CP.

## 2. Methods

This study was reviewed and authorized by the University of Minnesota institutional review board review (STUDY00012420). All experiments were performed in accordance with relevant guidelines and regulations. Informed consent for use of medical records was obtained at the time of service from all participants or their legal guardian. An option to rescind this permission is offered to patients at every visit to our center.

We built outcome prediction models that either withheld or included measurements only obtainable through IGA. We then tested the accuracy, responsiveness, and precision of these models. If the models including the IGA performed better than those without, then we concluded that IGA has value for the treatment of patients and that the use of ML prediction models is a sensible Step Two between collecting data and assigning treatments.

The models are of the form *y* = *f*(*X*), where *y* is an outcome of interest, and *X* are features drawn from clinical analysis (CLI) or from IGA. Note that these features (described below, details in Appendix 1) include interval treatments. In *training* the models, the interval treatments are the actual treatments administered and registered in our database. In *using* the models, the interval treatments would be proposed by the surgeon. Thus, once a model is built, it is possible to simulate the outcomes from a variety of different treatment combinations.

The model features consist of data elements commonly used in analyzing patients with mobility impairments. Some of these can be obtained using conventional means such as history and physical exam (*X*_CLI_) while some others are only derivable from an IGA (*X*_IGA_). The *X*_IGA_ are comprised of lower-limb joint angles, moments, power, and surface electromyographic (EMG) signals. Other IGA data elements (e.g., plantar pressure, walking energetics) are not included at this time due to their sporadic use in clinical gait laboratories. However, the models can be extended to include whatever CLI or IGA variables are available.

### Participants

We evaluated participants whose data was in our clinical database and who underwent IGA between January, 1994 and September, 2023. Our search criteria were as follows:

- Diagnosis of cerebral palsy (CP).
- Baseline and follow-up data available, where follow-up occurred within 2.5 years of baseline.
- Lower-limb joint angles present in baseline and follow-up data (note: moments, power, EMG not required).
- Age at baseline ≤ 18.

We randomly split the data into training and independent test sets (70%, 30%). Splitting was done on a by-visit basis. That is, no visits from the training set appeared in the test set. Because some patients had multiple visits, there were patients whose data appeared in both training and test data. This reflects a realistic application of the model: the model is trained at some point in time, this training includes data from *patient X*. Years later, *patient X* returns and the model is used for their current evaluation. We will show that the model performed similarly on previously unseen patients and those whose data appeared in the training data.

### Model Features

Features included in the CLI and IGA models are listed below (see Appendix 1 for a detailed list).

### Clinical Model (CLI)

This model only includes features available through a non-instrumented gait evaluation. For conciseness, we refer to this as the “CLI” model, rather than the “CLI - only” model.

- Comprehensive physical examination (PE) data including range-of-motion (ROM), bony torsion, strength, spasticity, static motor control, and foot structure.
- Interval treatment (between baseline and follow-up) comprised of 12 common surgeries accounting for 94% of all orthopedic surgery administered to the limbs included in this study. Adductor Release, Psoas Release, Hamstrings Lengthening, Calf Muscle Lengthening, Femoral Derotation Osteotomy, Tibial Derotation Osteotomy, Distal Femoral Extension Osteotomy (DFEO), Patellar Advancement, Foot and Ankle Bone Surgery, Foot and Ankle Soft Tissue Surgery, Selective Dorsal Rhizotomy, and Rectus Femoris Transfer.
- Birth history (weeks of gestation, time in NICU)
- Developmental history (age of walking onset)
- Current physical therapy (yes/no, frequency, location)
- Overall functional level (GMFCS, FAQ, FAQT) [14,15]
- Prior treatment history (yes or no) for the 12 common surgeries listed above

### Instrumented Gait Analysis (IGA) Model

This model includes features available through *both* non-instrumented and instrumented gait evaluation. We refer to this as the “IGA” model rather than the “IGA + CLI” model.

- All features from the CLI model
- Kinematics: lower-limb joint angle statistics (e.g., mean stance value, maximum swing value, …) and overall gait deviations as measured by the Gait Deviation Index (GDI) [16]
- Kinetics: lower-limb joint moment and power statistics
- Overall walking dynamic motor control as measured by the Walking Dynamic Motor Control Index (walk-DMC) [17]

### Demonstration

We will demonstrate our approach in detail using a pair of surgeries and a single outcome. The surgeries we highlight are femoral and tibial derotation osteotomies (FDO and TDO) and the outcome is mean stance-phase foot progression. We choose these two treatments and this outcome for the following reasons:

- Excessive in- and out-toeing are common in CP and impact both gait biomechanics and patient well-being (body image and self-esteem) [18].
- Both FDO and TDO are commonly administered at our center (see Figure 3 below).
- Both FDO and TDO have large effects at the structural level [3].
- A previous analysis identified a sub-group of limbs undergoing FDO who exhibited poor outcomes, seemingly due to sub-optimal treatment decisions. Specifically, some patients exhibited excessive external foot progression following an FDO. These limbs presented at baseline with excess anteversion but dynamic hip rotation during gait that was within normal limits [19].
- Change in foot progression is (primarily) affected by FDO, TDO, and foot surgery. This allows us to demonstrate the ability of the proposed approach to model combinations of surgeries.
- The previously described RCT examined the role of expert opinion, guided by gait analysis, on FDO and foot progression.

**Figure 3:**
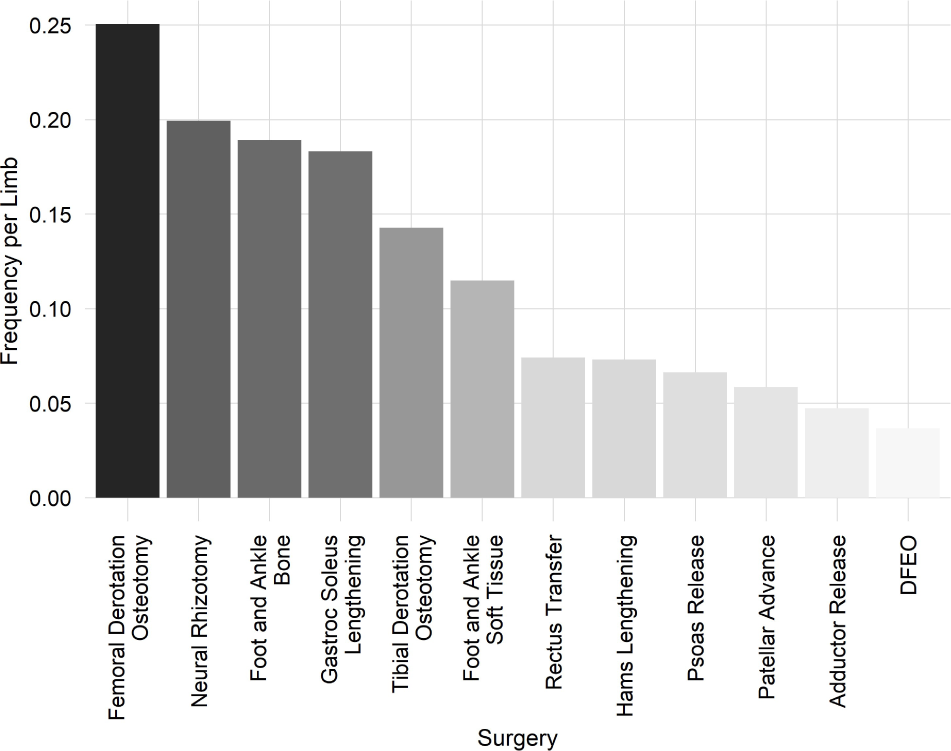
The frequency of surgical procedure per limb reflects the underlying treatment philosophy at our center plus the nature of patients referred for gait analysis.

Keep in mind that the outcome of choice is flexible. Here we have chosen the pre-to-post change in an angle, but we could have chosen the follow-up value of the angle, or another variable altogether (e.g., speed, GDI, etc.). It is merely a matter of inserting an alternative estimand into the ML model. Of course, not all estimands will be predicted with equal accuracy, precision, and responsiveness for all treatment combinations — but that will be reflected in the reported uncertainty. We will explore this issue in our results.

### Model Details

We use Bayesian Additive Regression Trees (BART) to predict mean stance foot progression after FDO and TDO. The estimation models are built using the BartMachine package in R with default parameters [20,21]. Many modeling frameworks can be used — from linear regression to neural networks. We chose BART for a number of reasons:

- BART generally requires no tuning [22]. In this study we use default parameters - though hyperparameter optimization is straightforward.
- BART is well regularized by default due to its Bayesian underpinnings (this is related to the above point regarding tuning).
- BART has been shown to perform exceptionally well in real and synthetic datasets, and this has been attributed to its ability to model complex response surfaces [23,24].
- BART produces an explicit, nonparametric distribution of possible outcomes (Bayesian Posterior). This can be used to determine probabilities of reaching various outcome thresholds.
- BART handles missing data seamlessly, without the need for imputation [25]. Note that missing data is allowed in all features.
- BART is fast and simple to use.

### Model Performance

We evaluated the accuracy of models by the root mean squared error (RMSE). We evaluated the ability of models to capture heterogeneous treatment effects (responsiveness) using the standard goodness-of-fit measure (R^2^) for the measured outcome as a linear function of the predicted outcome. We evaluated the precision of the models by the 90% and 50% Bayesian posterior *prediction* intervals.

## 3. Results

### Limb Characteristics

There were 5834 limbs returned by the database query (Table 1). This represented both limbs from 1556 patients seen over 2917 separate examinations. The large number of limbs with missing DMC values is due to the fact that our center did not routinely collect post-treatment EMG until around 2012. In what follows, all measures of model performance are based on the independent test set.

**Table 1:** Limb Characteristics.

| Characteristic | Train, N = 4,084<sup>1</sup> | Test, N = 1,750<sup>1</sup> |
| --- | --- | --- |
| CP Subtype |  |  |
| Diplegia | 2,488 (61%) | 1,140 (65%) |
| Hemiplegia | 184 (4.5%) | 72 (4.1%) |
| Hemiplegia type I | 14 (0.3%) | 4 (0.2%) |
| Hemiplegia type II | 80 (2.0%) | 40 (2.3%) |
| Hemiplegia type III | 38 (0.9%) | 18 (1.0%) |
| Hemiplegia type IV | 56 (1.4%) | 14 (0.8%) |
| Quadriplegia | 560 (14%) | 202 (12%) |
| Triplegia | 660 (16%) | 258 (15%) |
| Missing | 4 (<0.1%) | 2 (0.1%) |
| Age [yr] | 8.2 (6.0, 11.2) | 8.3 (6.2, 11.6) |
| Sex |  |  |
| F | 1,666 (41%) | 750 (43%) |
| M | 2,418 (59%) | 1,000 (57%) |
| GMFCS |  |  |
| I | 1,162 (28%) | 504 (29%) |
| II | 1,594 (39%) | 692 (40%) |
| III | 1,220 (30%) | 506 (29%) |
| IV | 78 (1.9%) | 32 (1.8%) |
| Missing | 30 (0.7%) | 16 (0.9%) |
| GDI | 69 (61, 76) | 69 (60, 76) |
| walk-DMC | 80 (73, 86) | 79 (72, 86) |
| Unknown | 1,852 | 774 |

<sup>1</sup>n (%); Median (IQR)

### Treatments

Limbs in the dataset underwent a variety of treatments reflecting both the type of patient referred to our center, as well as the local treatment philosophy. We can examine the distribution of 12 common surgeries in this cohort (Figure 3). Note that this list excludes several infrequent or minimally impactful treatments, including neurolytic injections (e.g., botulinum toxin type A), casting, spine surgery, total knee arthroplasty, limb lengthening, baclofen pump implantation, instrumentation removal, casting, upper extremity surgery, and exploratory surgery.

We will present a detailed examination of limbs that underwent an FDO, a TDO or both. Generally, these surgeries are not administered in isolation. We can examine the distribution of concomitant surgeries in limbs that underwent an FDO or a TDO (Figure 4).

**Figure 4:**
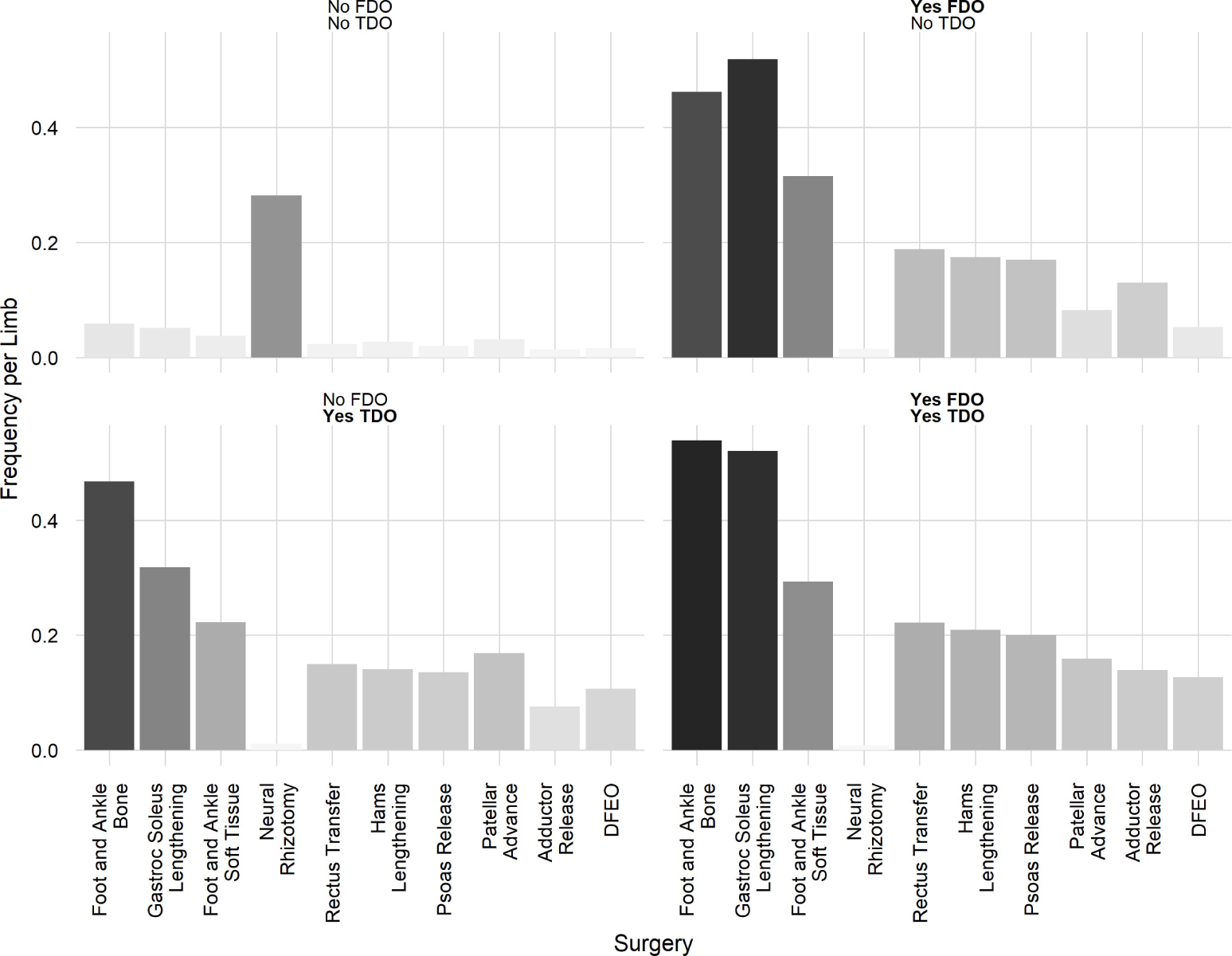
Concomitant treatment for limbs undergoing FDO or TDO.

### Model Performance

Overall, the IGA model was more accurate than the CLI model (RMSE_IGA_ = 11^∘^, RMSE_CLI_ = 13^∘^) and explained almost twice as much of the variance in the measured outcome (R^2^_IGA_ = 0.45, R^2^_CLI_ = 0.28) (Figure 5). The IGA model calibration data were evenly dispersed, while the clinical model exhibited a non-uniform distribution, with a large percentage of predictions clustered around zero. Both models appear unbiased, and have a calibration slope of around one.

**Figure 5:**
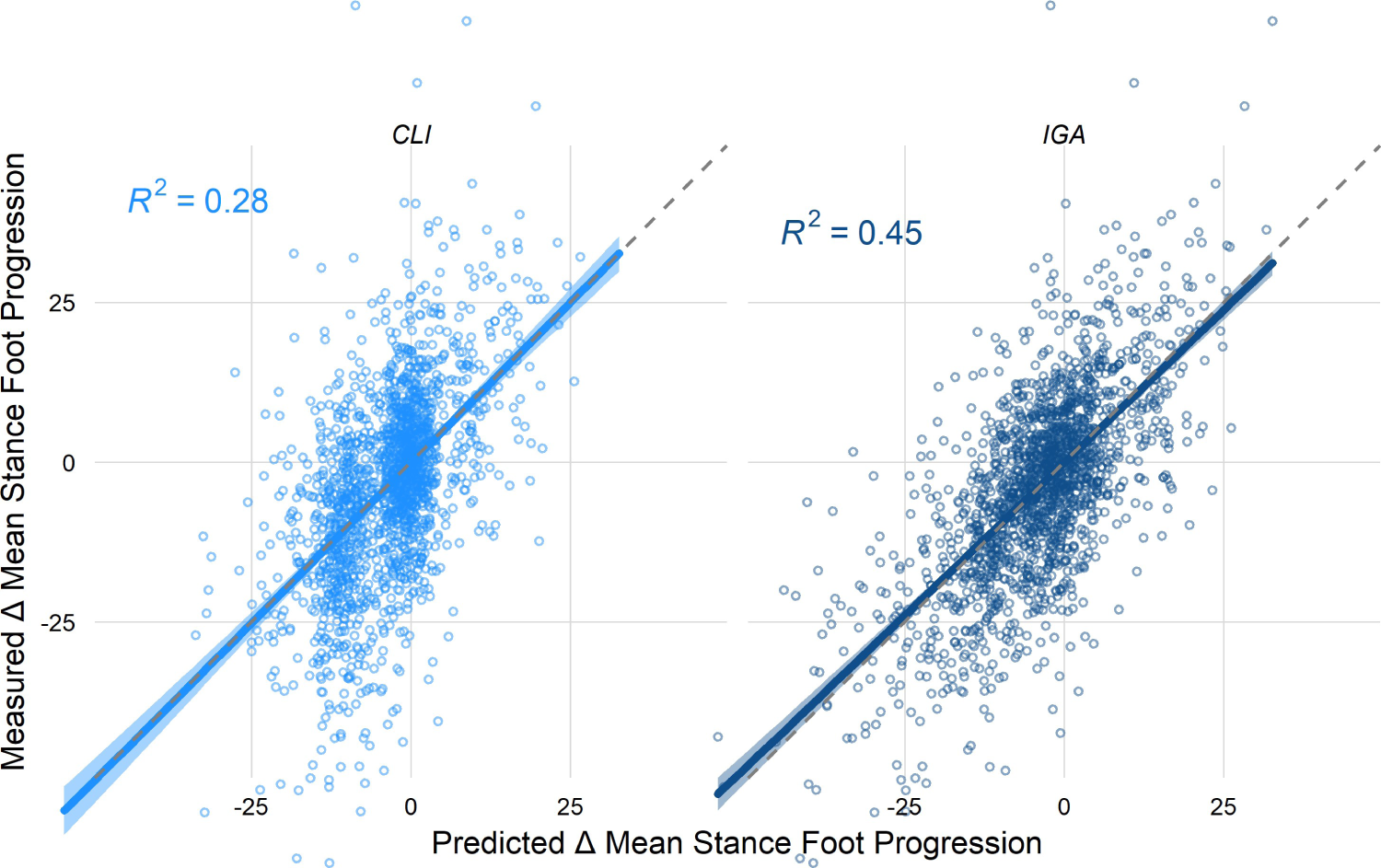
Overall performance of the IGA and CLI models.

We next stratified the results by whether or not a limb underwent an FDO or a TDO. Note that this analysis averages over other treatments (e.g., bony foot surgery, gastrocnemius lengthening) that can affect foot progression. However, FDO and TDO have large impact on foot progression, and the concomitant treatments are similarly distributed among these strata (see Figure 4), so we will focus on these to demonstrate the models’ performance. This simplification would not be necessary when using the model in a clinical setting, since the status of other treatments can be assigned explicitly.

Visually, the overall behavior of the models across the four treatment options Yes/No FDO ∩ Yes/No TDO suggest the superiority of the IGA model (Figure 6). The IGA model fits the measured data better, and appears more accurate (quantified below).

**Figure 6:**
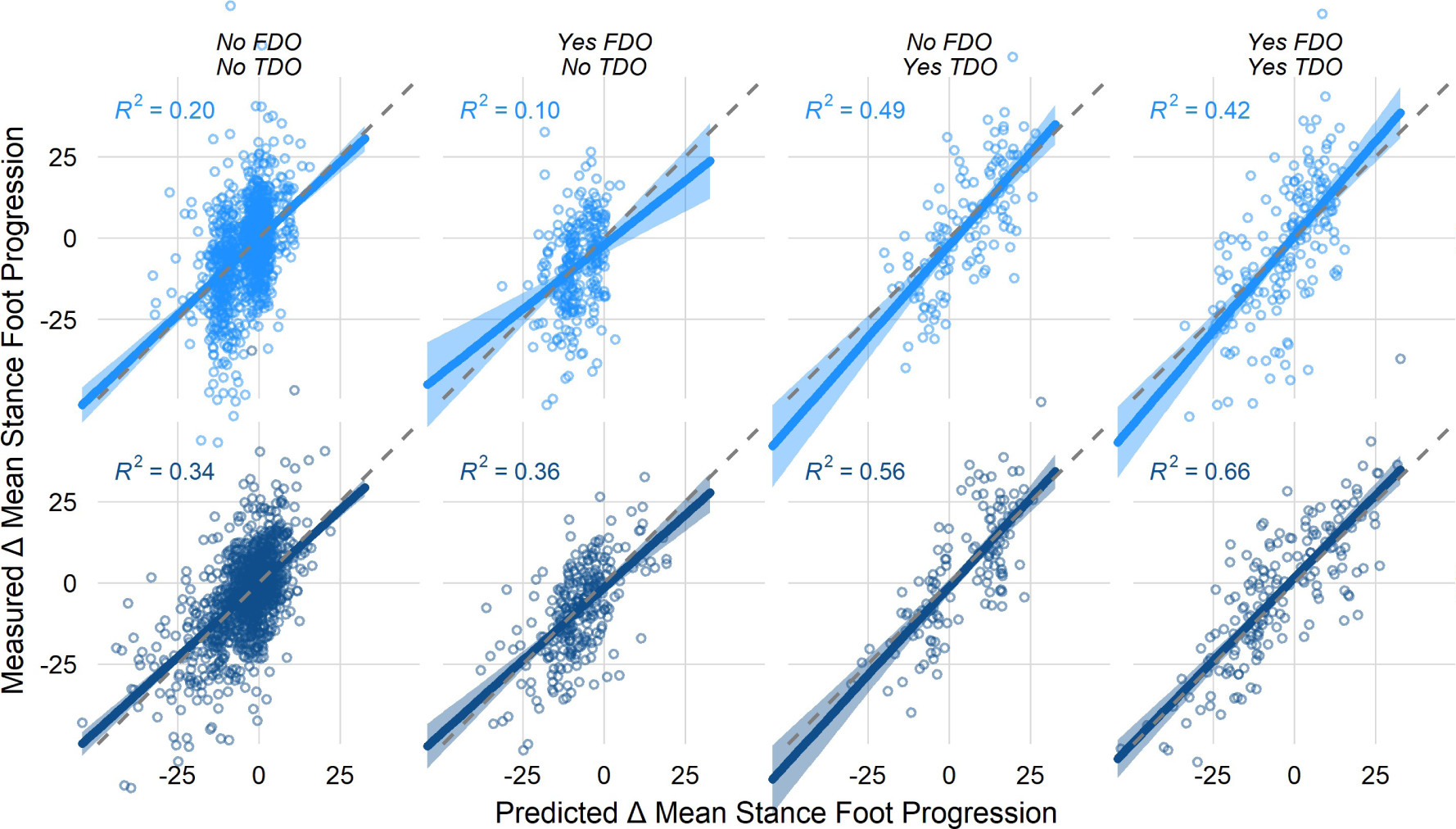
Comparison of CLI (top) and IGA (bottom) models. The predicted (x-axis) and measured (y-axis) results are shown for mean stance-phase foot progression angle. Columns reflect FDO and TDO status. Visually, it is apparent that the IGA model is more accurate and captures a wider range of treatment responses. These subjective impressions will be quantified below.

The IGA model had a lower RMSE and a higher R^2^ across every treatment subgroup (Figure 7). The accuracy superiority ranged from 1^∘^ - 4.5^∘^ RMSE. In terms of capturing outcome heterogeneity, the IGA model was substantially better — explaining from 1.5 to more than 2 times the variance in the outcome compared to the CLI model.

**Figure 7:**
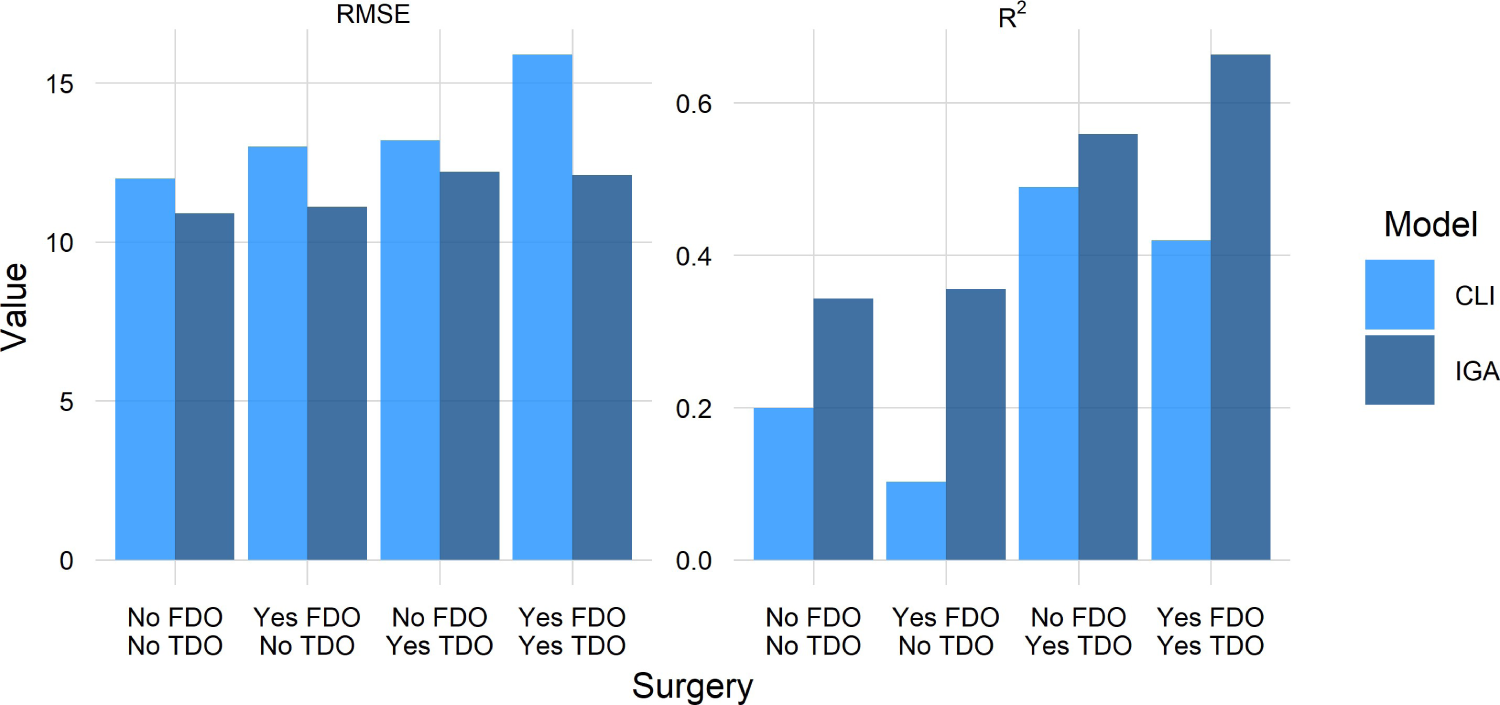
RMSE lower (good) and r-squared higher (good) for IGA.

It is reasonable to wonder how well a prediction model performs with increasing treatment complexity. It would be conceivable that a model would work well for untreated or sparingly treated limbs, but would fail for limbs undergoing many surgeries. It is common that children with CP undergo many surgical procedures simultaneously. Planning these complex treatments was part of the original rationale for IGA. As the number of simultaneous surgeries increased, the IGA and CLI models became slightly less accurate, with the IGA model performing better at every level (Figure 8). At 6+ simultaneous surgeries the IGA RMSE only increased slightly while the CLI model degraded substantially. The IGA model demonstrated a large advantage in R^2^, especially at the complexity extremes.

**Figure 8:**
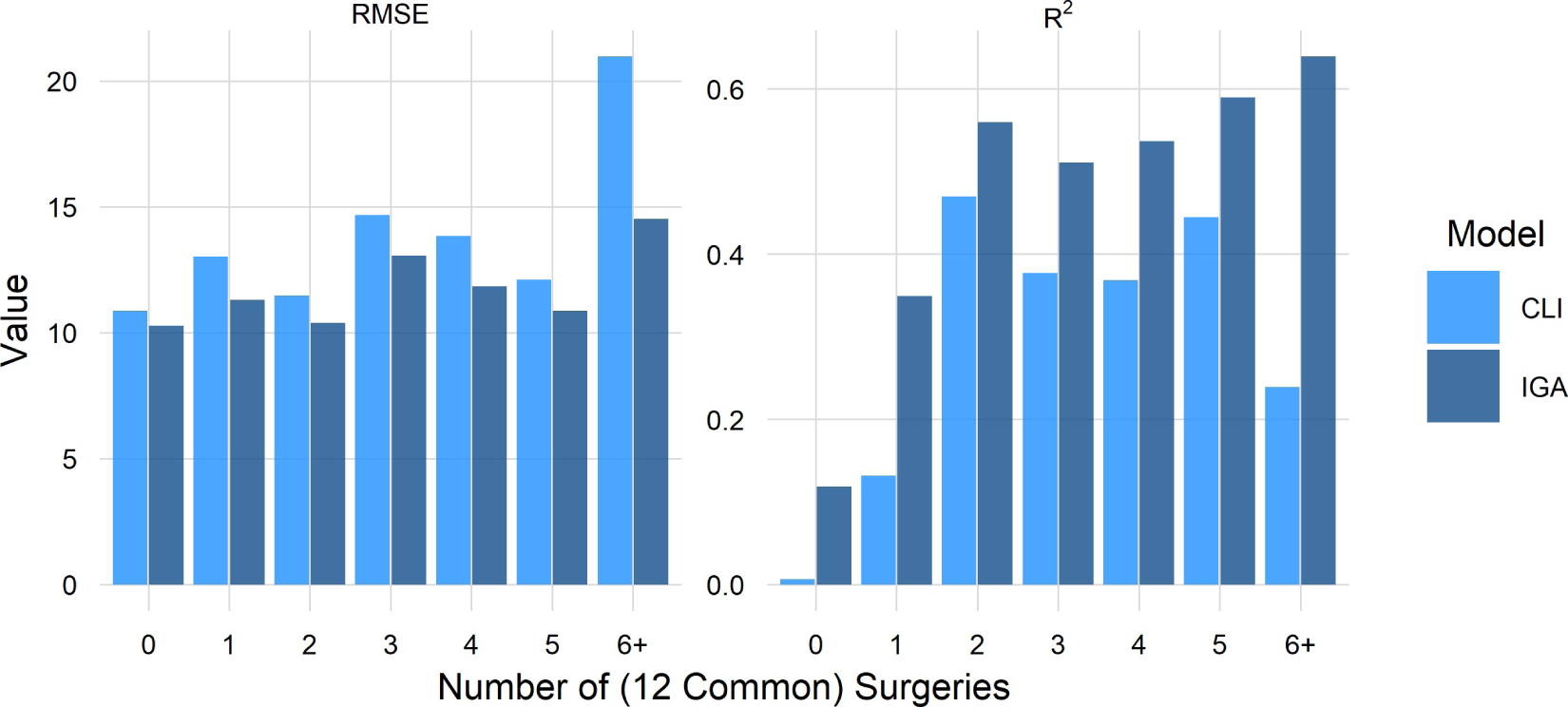
IGA model does better at every level of complexity.

Some patient limbs appeared in both the training and test data — though, as described above, not from the same visit. We tested whether this introduced any optimistic bias to the results by examining the accuracy (RMSE) on limbs from never-before-seen patients compared to those from patients who appeared in the training data (Figure 9). We found no meaningful bias, suggesting future performance should reflect the results presented here.

**Figure 9:**
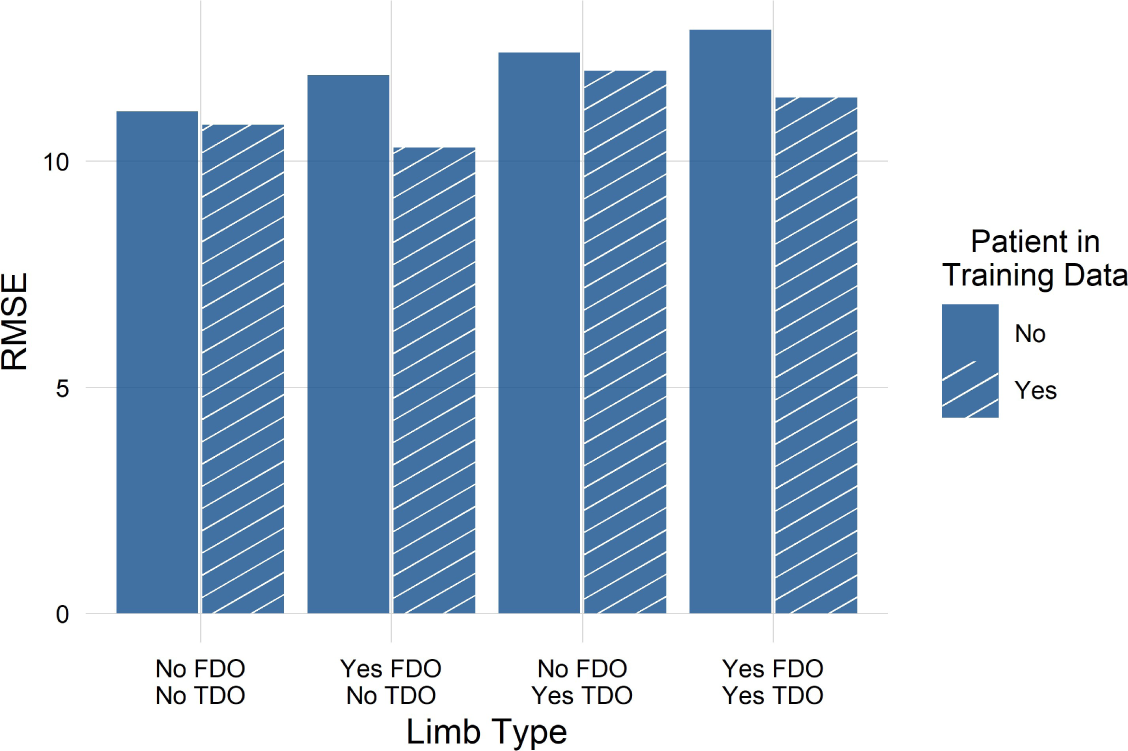
No evidence of meaningful optimistic bias in the IGA model. In fact, for some categories of limbs the repeated data exhibited slightly lower accuracy.

For all categories of limbs Yes/No FDO ∩ Yes/No TDO, the IGA model was more precise than the CLI model (Figure 10). The 90% *prediction* intervals for the IGA model were around 6^∘^ narrower than those for the CLI model. The differences for the 50% CIs were around 2.5^∘^.

**Figure 10:**
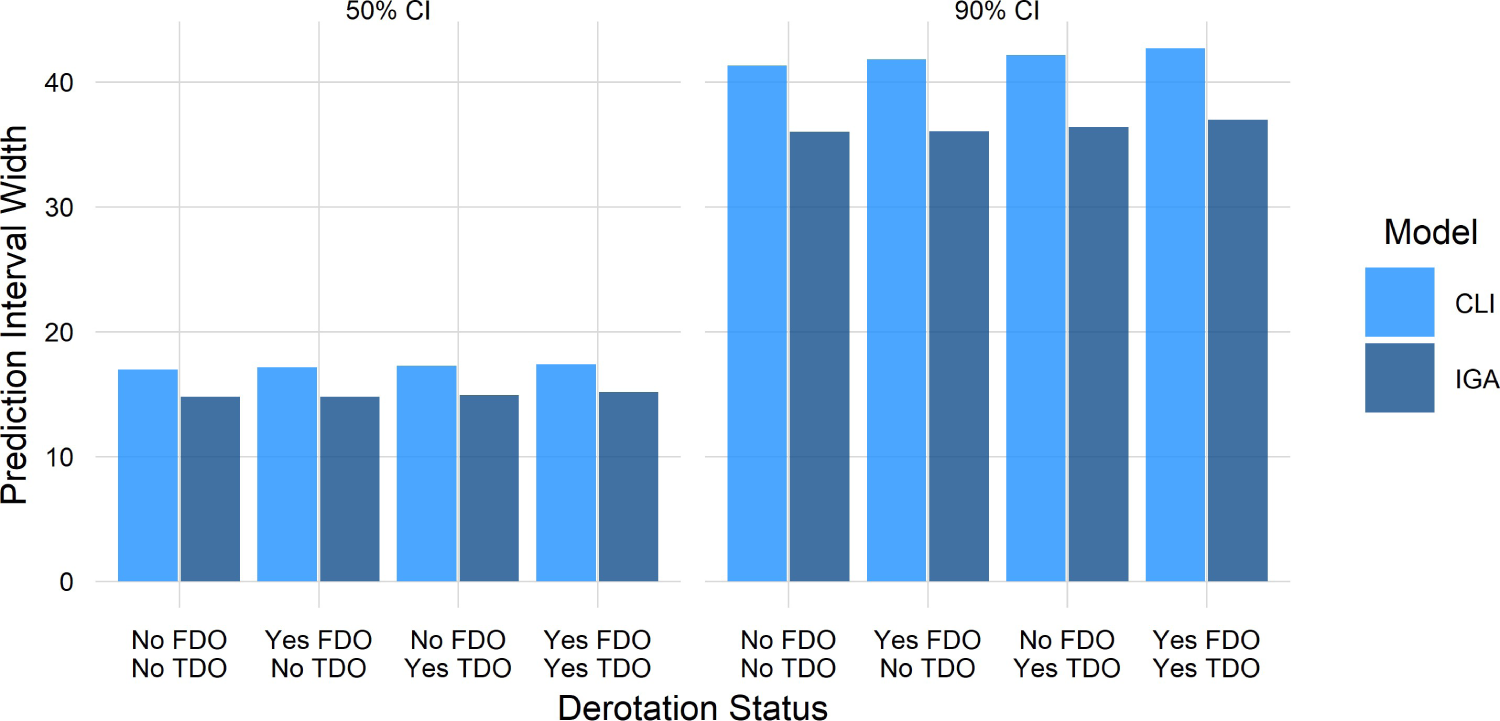
Precision (CI) of CLI and IGA models. A smaller value reflects a more precise prediction, and thus indicates superior performance.

Note that the 90% prediction intervals are wide — around 35^∘^. Keep in mind however, that precisely predicting a particular patient’s post-op predicament for any procedure is preposterous. Even extremely common and highly studied procedures, such as treatment of blood pressure, total joint replacement, and so on, have sizeable outcome uncertainty for individuals.

To put these prediction intervals in perspective, in current practice the uncertainty bounds for *individualized* outcome prediction are NaN. That is, they are undefined. They don’t exist. There’s nothing — it is just Kentucky windage. Clinicians and patients currently have no explicit outcome estimate available to them. Any vague estimates that do exist are *group* response means and standard deviations (i.e., average treatment effects) from studies whose participants may be completely dissimilar to “*the patient in front of you*”.

An important point to appreciate is that any prediction interval — not just 90% — can be computed from the ML model. The computed outcome probability can be tailored to the patient’s risk tolerance and the potential benefits of the treatment. The predictions can also be “thresholded” to provide the probability of attaining a certain outcome level. For example, “*how likely is it that foot progression will improve by more than 10*^∘^”.

### Case Study

We present a case study showing the utility of the model to demonstrate how this approach might look in practice. We examine a limb from a typical patient with torsional deformities who underwent an FDO and TDO, along with several other surgeries.

### Baseline

The limb was from a 6-10 year old male diagnosed with the Diplegia subtype of CP who functioned at a GMFCS level I. At baseline, the limb’s transverse plane profile indicated excessive femoral anteversion, external tibial torsion (Table 2). Dynamically, there was internal rotation of the hip, external rotation of the knee, and internal foot progression during gait (average during the stance-phase of gait).

**Table 2:**
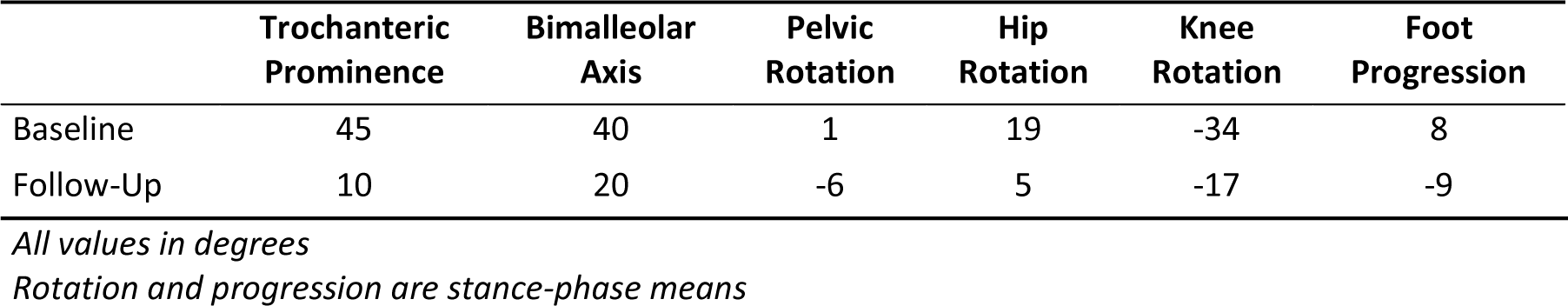
Torsion Profile.

|  | **Trochanteric Prominence** | **Bimalleolar Axis** | **Pelvic Rotation** | **Hip Rotation** | **Knee Rotation** | **Foot Progression** |
| --- | --- | --- | --- | --- | --- | --- |
| Baseline | 45 | 40 | 1 | 19 | -34 | 8 |
| Follow-Up | 10 | 20 | -6 | 5 | -17 | -9 |

*All values in degrees*
*Rotation and progression are stance-phase means*

### Treatment

The limb underwent the following treatments: Femoral Derotation Osteotomy, Foot and Ankle Soft Tissue, Patellar Advance, Tibial Derotation Osteotomy.

### Follow-Up

The patient was followed-up approximately 1 year later. The measured change in foot progression was −17^∘^. The IGA model predicted a change in foot progression of −15^∘^ with a 50% prediction interval of −23^∘^ to −7^∘^.

The trochanteric prominence test angle changed by −35^∘^ and the bimalleolar axis angle changed by −20^∘^. The IGA data showed mean stance phase transverse plane pelvic, hip, knee, and foot angle changes of −7^∘^, −14^∘^, 17^∘^, and −17^∘^, respectively. These are consistent with an externalizing femoral osteotomy and an internalizing tibial osteotomy. Note that the change in foot progression does not necessarily equal the summed derotation magnitudes. The impact of derotations on transverse plane kinematics is known to not be one-to-one [18,26].

### Counterfactual Treatments

To understand the impact of treatment it is important to compare the effect of treatment to the untreated natural history. We can do this by setting all interval treatments to FALSE. For the limb in question, the predicted short-term untreated change in foot progression and 50% prediction interval was −7^∘^ (−14^∘^, 0^∘^). This change can be plausibly attributed to remodeling of bony torsion, or other changes in gait pattern that occur with maturation, given the patients relatively young age.

We could also predict the outcome for any combination of surgeries. Many combinations would not be clinically sensible. However, *if* the purpose of the derotations was to externalize foot progression, it is of interest to estimate what outcome could be achieved with an isolated derotation of the femur. To simulate the alternative prescription, we set TDO to FALSE, leaving all other surgeries as is. The model predicted a change in foot progression and 50% prediction interval of −13^∘^ (−21^∘^, −6^∘^). Note that since the tibia already has an external deformity, it would not be sensible to administer an externalizing derotation osteotomy at that level. Also keep in mind that a TDO and FDO may be administered in tandem to address knee pain often attributed to the combined malalignment of excessive anteversion and external tibial torsion.

Lastly, we might be interested in whether any of the other surgeries contributed to the change in foot progression. We estimate this by setting FDO and TDO to FALSE, leaving all other treatments as-is, and predicting the outcome. We found the predicted outcome almost equals the no-treatment case described above (−7^∘^ vs. −7^∘^, for no FDO/TDO vs. no treatment). This suggests that, for this patient, the model does not ascribe substantial foot progression changing capacity to treatments other than derotational osteotomies. This is sensible, given the list of other treatments (Foot and Ankle Soft Tissue, Patellar Advance).

### Other Outcome Measures

It is logical that IGA would be most useful for predicting gait outcomes, or outcomes that are directly affected by gait. We examined overall model performance for six additional measures. Three of these were specific gait parameters related to crouch (mean stance knee flexion), equinus (mean stance ankle dorsiflexion) and stiff-knee gait (maximum swing knee flexion). These are common impairments that are frequently addressed with surgery. Two other outcomes were related to overall gait pattern (GDI and dimensionless speed), while the final was overall mobility (FAQT), which is indirectly affected by gait. Note that the first three outcomes are relatively responsive to treatment and are directly related to gait.

We expect IGA to be valuable in these situations. The next three outcomes (GDI, speed, and FAQT) are related to gait, but are relatively or entirely non-responsive to treatment, and are strongly influenced by untreated factors such as poor motor control and weakness.

Examining the accuracy of the models, and their ability to explain heterogeneous outcomes, we see an advantage for the IGA model (RMSE and R^2^) in every measure except mobility (FAQT) (Figure 11). As expected, IGA adds more value for responsive variables directly related to gait.

**Figure 11:**
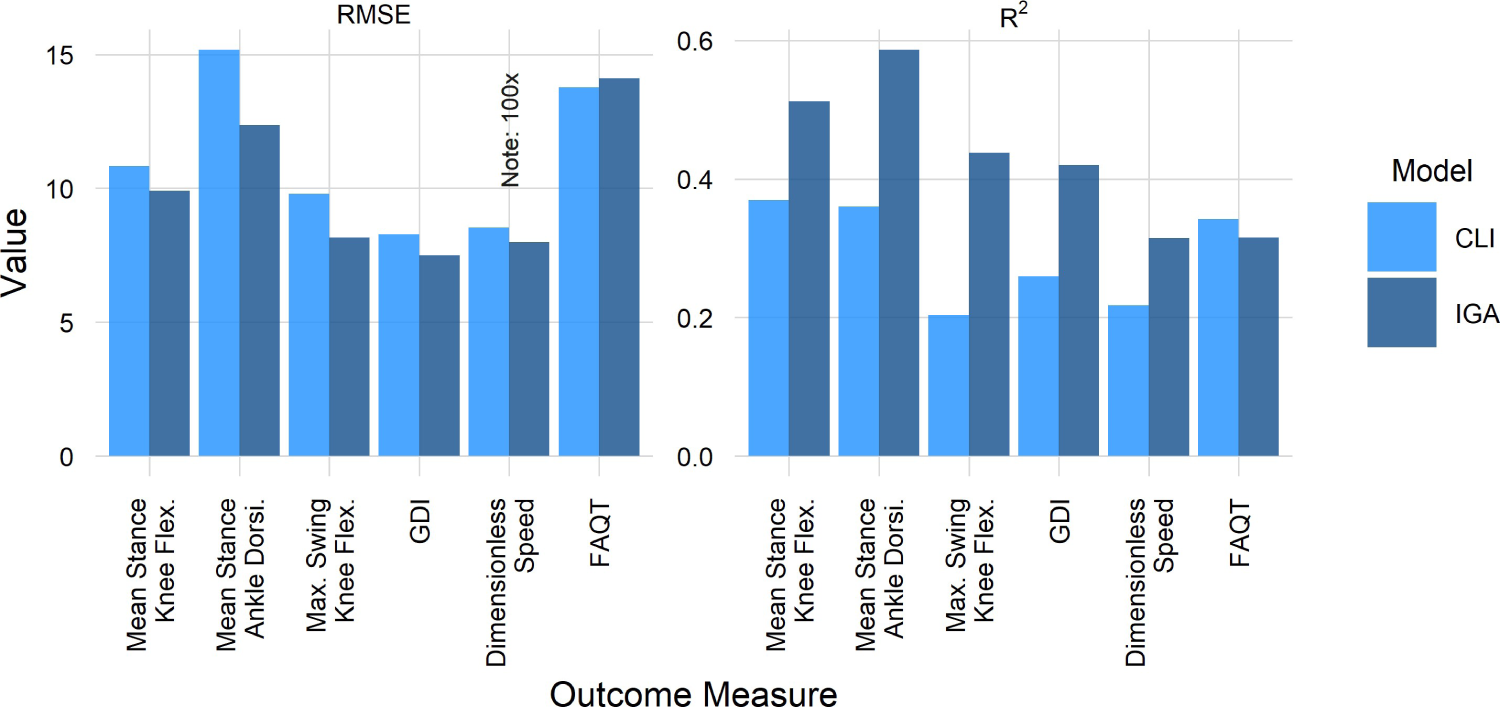
Better (almost) everywhere. Note speed RMSE plotted at 100x to put it on similar scale as other outcomes.

For variables that are causally upstream from gait (e.g., femoral anteversion), we do not expect — or find — IGA to add value to outcome prediction (RMSE/R^2^ CLI = 12.2^∘^/0.71, RMSE/R^2^ IGA = 12.3^∘^/0.7).

## 4. Discussion

We demonstrated that IGA has clear *potential* value in the treatment of mobility impairments common to children diagnosed with CP. The addition of data from IGA allows ML models to predict treatment outcome with greater accuracy, precision, and responsiveness compared to using CLI data alone. However, realizing this potential requires a fundamental change in how IGA data is used.

Our demonstration of IGA’s potential requires us to employ outcome estimation models as part of the treatment assignment process. It is simply not possible for ordinary human minds to synthesize the massive amount of data a typical IGA produces, while simultaneously incorporating the historical outcomes, individual risk factors, uncertainty in various data elements, correlations among the multidimensional data, and so on.

Our recommendation is to replace a significant portion of the current standard approach — the subjective “*meeting of the minds*” described earlier — with explicit outcome predictions. These predictions would be used along with — not instead of — the usual clinical impressions divined from history, video, knowledge of the patient, and so on. It is important to be honest about the fact that much of what determines the patient outcome (unmeasured clinical features, surgical skill, postoperative care, patient motivation, family resources, etc…) is not captured by these models. They only reflect — in an objective, data-driven manner — the information obtainable from a comprehensive gait evaluation. In this regard, the ML outcome models form a valuable decision support scaffolding for, not a replacement of, the existing clinical process.

It would be interesting to investigate which IGA data elements are most useful. For example, are kinetic data (joint moments and power) worthwhile? Does the EMG data add value? What about plantar pressures, walking energetics, …? We can ask the same question about the clinical data. Are there elements of the physical exam that are not necessary? The answers to these questions can allow us to streamline the IGA process. Currently, a comprehensive IGA evaluation at our center takes about 2.5 hours. Reducing the evaluation to those elements essential for treatment assignment can save time, money, and make the ML-aided interpretation simpler and better targeted to patient goals.

Another obvious extension is examine which outcomes are best estimated using IGA. Can we predict quality of life? What about pain? Prior causal analyses suggest only a weak connection between gait and many “*macro*” scale outcomes like mobility. Answering this scope question would allow us to tailor gait analysis and its application to domains in which it has relevance.

### Step Two — The Devil is in the Details

We have simultaneously demonstrated the utility of IGA and proposed a scheme for harnessing that utility via ML outcome estimation models. But how do we get from the current IGA paradigm to what is proposed here?

An obvious challenge is how to communicate this type of information to both clinicians and patients. This is a known obstacle in the real-world use of ML modeling. Communication of complex data is a robust and growing field, and we are confident that there are ways to overcome this challenge (e.g., [27]). We are currently experimenting with several user interfaces and outcome formats. The interfaces would allow clinicians and patients to easily enter the outcomes that are of interest to them and the treatments that are reasonable, based on historical data. The output is a compact summary showing the probability of reaching the specified outcome goal.

We carefully examined a single outcome in response to two possible treatments. These are common treatments in our database, and an outcome that tends to respond robustly. We also showed that several other outcomes behaved similarly — though IGA added the most value when the outcomes were gait-related and responsive to treatment. We have previously shown that treatment effects are attenuated as they travel through the causal chain [3]. Likewise, we expect the value of IGA to be smaller and less clear as we move further down the causal chain from gait → mobility → participation → quality of life.

The treatments we focused on — FDO and TDO — are two of the most common treatments in our database and have fairly clear indications. Less common treatments, those with smaller impacts, and treatments with less well-agreed on indications (e.g., psoas lengthening) may be harder to model. In these cases, however, there is a built-in safety mechanism. Namely, the modeling framework we use is honest and conservative. If there is no strong evidence to predict an outcome, then the prediction will have wide uncertainty bounds. The reporting of uncertainty in a way that is properly appreciated and understood is absolutely critical to the proper and ethical use of any modeling framework.

### Strengths and Limitations

As stated at the outset, the proposed version of Step Two is evidence-based, transparent, and objective. As such it provides a sensible way for using gait data to support treatment decisions. It can be clearly documented in patients’ medical record and allows the potential benefits of various treatments to be explicitly explained by the treating clinician. The nature of the proposed Step Two also allows the framework to be periodically tested for value, and to be updated — or abandoned — if it performs below expectations.

The current models include common data obtained from an IGA evaluation. The approach is entirely flexible. Modeling new outcomes is simply a matter of swapping in a new estimand — as we did in the “Other Outcomes” analysis — while the addition of new predictors is only a matter of adding them to the feature set *X*. For example, our center routinely acquires image-based estimates of various structural measurements like femoral anteversion and tibial torsion. Adding these as predictive features may improve the accuracy, responsiveness, or precision of the models. It is also possible to add treatment dosing details. For example, instead of simply indicating a limb will undergo an FDO, the proposed amount of derotation could be included. We have run initial tests with both of these enhancements and observed small improvements to model performance.

While many modeling frameworks can be used (e.g., linear regression, neural networks), we find the BART approach works well. In particular, BART handles missing data seamlessly, without the need for subjective imputation algorithms. Missing data is common in the patient population we are considering, and is often *not* missing at random. For example, children with greater degrees of impairment are often unable to complete parts of the clinical evaluation (e.g., missing strength and motor control testing due to inability to understand and follow directions) or parts of the instrumented gait analysis (e.g., missing kinetics due to use of a walker or crutches). A Step Two framework that is limited by these constraints would be less useful. Importantly, it would also be less just, since it would systematically exclude a particularly vulnerable portion of the population that it is intended to serve.

Not every laboratory has an extensive database and in-house machine learning expertise. We are sharing the foot progression model and a sample dataset for other researchers to use, test, and improve. Caution is necessary, as we have not established generalizability of models built on our data to other centers. There is an ongoing project that will pool clinical gait data from a large number of centers and evaluate outcome prediction models like those described here [28]. The results of this large study will provide strong evidence for or against the use of outcome estimation models as a clinical tool.

## Conclusion

> “Some people laughed to see the alteration in him, but he let them laugh, and little heeded them; for he was wise enough to know that nothing ever happened on this globe, for good, at which some people did not have their fill of laughter in the outset”

> — Charles Dickens, A Christmas Carol

To date, clinical gait analysis has not been shown to improve outcomes for children with mobility impairments. If IGA continues its present course, then its value will remain unrealized, and outcomes will remain modest, stagnant, and unpredictable. In this manuscript we have described how IGA can be used to generate explicit estimates of treatment outcomes. By using these estimates, alongside the existing array of standard clinical practices, IGA can become as good a technology, as good a clinical tool, and as good an estimator of outcome, as the good old surgeon knows, or any other good old clinician, specialist, or sub-specialist, in the good old world.

## Data Availability

All data produced in the present study are available upon reasonable request to the authors

## Appendix 1: Model Features

### Clinical Features

Features for the CLI model are drawn from the standard clinical evaluation that includes patient history and physical examination. The names listed below are codes from our database, but most of them are intelligible.

### Diagnosis

dxmod, affected

### Descriptive

age, HEIGHT, WEIGHT, Sex, SIDE

### Birth and Developmental History

NICU_Weeks, Delivery_Weeks, Ventilator_Weeks, AGE_AT_DIAG, DEV_FIRST_STEP, DEV_WALK

### Function and Mobility

GMFCS, FAQ, FAQT

### Prior and Interval Treatment

prior_Adductor_Release, prior_Psoas_Release, prior_Tibial_Derotation_Osteotomy, prior_Foot_and_Ankle_Bone, prior_Gastroc_Soleus_Lengthening, prior_Femoral_Derotation_Osteotomy, prior_Hams_Lengthening, prior_Rectus_Transfer, prior_Foot_and_Ankle_Soft_Tissue, prior_DFEO, prior_Patellar_Advance, prior_Neural_Rhizotomy, interval_Adductor_Release, interval_DFEO, interval_Femoral_Derotation_Osteotomy, interval_Foot_and_Ankle_Bone, interval_Foot_and_Ankle_Soft_Tissue, interval_Gastroc_Soleus_Lengthening, interval_Hams_Lengthening, interval_Neural_Rhizotomy, interval_Patellar_Advance, interval_Psoas_Release, interval_Rectus_Transfer, interval_Tibial_Derotation_Osteotomy

### Range of Motion and Alignment

HIP_EXT, HIP_ABD_0, POP_ANG_UNI, KNEE_EXT, EXTEN_LAG, ANK_DORS_0, ANK_DORS_90, ANTEVERSION, BIMAL

### Strength, Spasticity, Static Motor Control

ABDOM_SEL, ABDOM_STR, BACK_EXT_SEL, BACK_EXT_STR, ADDUCTOR_SPAS, ANT_TIB_SEL, ANT_TIB_STR, EXT_HALL_LONG_SEL, EXT_HALL_LONG_STR, FLEX_HALL_LONG_SEL, FLEX_HALL_LONG_STR, HAMSTRING_SPAS, HIP_ABD_SEL, HIP_ABD_STR, HIP_ADD_SEL, HIP_ADD_STR, HIP_EXT_KN90_SEL, HIP_EXT_KN90_STR, HIP_EXT_SEL, HIP_EXT_STR, HIP_FLEX_SEL, HIP_FLEX_SPAS, HIP_FLEX_STR, KNEE_EXT_SEL, KNEE_EXT_STR, KNEE_FLEX_SEL, KNEE_FLEX_STR, PERON_BREV_SEL, PERON_BREV_STR, PERON_LONG_SEL, PERON_LONG_STR, PLANTFLEX_SEL, PLANTFLEX_SPAS, PLANTFLEX_STR, POST_TIB_SEL, POST_TIB_SPAS, POST_TIB_STR, RECT_FEM_SPAS

### Gait Features

We consider the following kinematic curves: Pelvis (all planes), Hip (all planes), Knee (all planes), Ankle (sagittal), Foot (transverse)

We consider the following moment curves: Hip (sagittal, coronal), Knee (sagittal, coronal), Ankle (sagittal)

We consider the following power curves: Hip, Knee, Ankle

We extract values (and timing) at the following instants: initial contact, opposite foot off, mid-stance, opposite foot contact, foot off, and mid-swing

We compute the following statistics (and timing where applicable): mean stance, maximum instance, minimum instance, mean swing, maximum in swing, minimum in swing

We also include the gait deviation index (GDI) and walking dynamic motor control (DMC)

